# Diastolic Perfusion Pressure Predicts Response to Inotropes/Vasopressors and Benefit from Mechanical Circulatory Support in Cardiogenic Shock?

**DOI:** 10.1101/2025.01.30.25321443

**Authors:** Hoong Sern Lim, Dagmar Vondrakova, Jan Belohlavek, Richard Rokyta, Petr Ostadal

## Abstract

**Background:** Hemodynamic response to escalation of vasoactive drugs has not been well-characterized in patients with cardiogenic shock CS. We tested the hypothesis that lower diastolic perfusion pressure (DPP=diastolic blood pressure-right atrial pressure) was associated with more limited hemodynamic response to up-titration of vasoactive drugs and with possible benefit from early mechanical circulatory support (MCS) in patients with CS.

**Methods:** This study consisted of two parts. (i) We evaluated the relationship between baseline DPP and changes in cardiac power output index (CPOI) in response to increase in vasoactive drugs in a cohort of patients with CS (n=93). (ii) We compared all-cause mortality based on baseline DPP in a post hoc analysis of the ECMO CS trial. CPOI responders were defined as post-escalation CPOI≥0.28 W/m^2^.

**Results:** Vasoactive inotrope score escalated from 11.2±3.9 to 24.5±4.7. Escalation of vasoactive drugs was associated with increases in CPOI to 0.23±0.06 W/m^2^ (all p<0.001).

Post-escalation CPOI was directly related to baseline cardiac index and DPP. Baseline DPP discriminated CPO responders from non-responders with optimal cutoff of 37mmHg. Patients with baseline DPP≥37mmHg had greater CPOI increase and lactate clearance. In the ECMO-CS trial, patients with DPP˂37mmHg had lower mortality (HR 0.37, 95% CI 0.14-0.97, P=0.044) with immediate VA ECMO compared to early conservative management, but no significant difference in the subgroup with DPP≥37mmHg.

**Conclusion:** Lower DPP was associated with more limited hemodynamic response to escalation of vasoactive drugs and potentially greater benefit from early VA-ECMO in CS.

## INTRODUCTION

Cardiogenic shock (CS) is a highly lethal condition. Deaths from cardiogenic shock are primarily attributable to refractory shock or cardiac causes, despite advances in mechanical circulatory support (MCS)^1^. The management of CS has been extensively reviewed and discussed^2^, but the dilemma that clinicians face when managing a deteriorating patient with CS remains unanswered. Central to the “clinician’s dilemma” is the decision to continue with up-titration of vasoactive drugs or escalation to MCS in a deteriorating patient with CS. Understanding the patient’s response to inotropes and vasopressors is central to addressing the “clinician’s dilemma”. For example, a patient with CS who is unlikely to achieve the necessary hemodynamic response to inotropes/ vasopressors may benefit from early escalation to MCS. In this regard, characterization of the CS phenotype may guide decision-making, as response to inotropes and vasopressors is dependent on the physiological phenotype^3^.

There is increasing interest in the phenotype of abnormal vasodilatation in CS, mostly defined by low systemic vascular resistance and/or vasopressor dose used^4^. Low systemic vascular resistance and high norepinephrine doses have been reported to be associated with increased risk of death in patients with CS^5^. However, there are limitations in the use of systemic vascular resistance not least the requirement for pulmonary artery catheterization. We recently described the physiologic basis for diastolic perfusion pressure (DPP = diastolic blood pressure (DBP) – right atrial pressure (RAP)) as a measure of abnormal vasodilatation [Supplementary Data], and further demonstrated that in the post-heart transplant setting, lower baseline DPP (but not systemic vascular resistance) predicted subsequent MCS use^6^. There are no studies on DPP in patients with CS.

In this study, we hypothesized that lower baseline DPP was associated with more limited hemodynamic response to escalation of vasoactive drugs in patients with CS and thus, may benefit from earlier MCS. To test this hypothesis, firstly, we studied the relationship between baseline DPP and hemodynamic response to up-titration of vasoactive drugs in a cohort of patients with CS; and secondly, we performed a post hoc analysis of the Extracorporeal Membrane Oxygenation in the Therapy of Cardiogenic Shock (ECMO-CS) Trial to compare the outcomes of patients with CS supported by VA ECMO based on baseline DPP.

## METHODS

### The Cardiogenic Shock Cohort

The CS cohort included 93 consecutive patients with CS from April 2016-April 2024 from a single centre. Patients who were deteriorating rapidly and too unstable for pulmonary artery catheterization (n=12), ongoing cardiac arrest and tamponade were excluded. Right heart catheterization was performed in the standard manner with a balloon-tipped, fluid-filled 7.5F Swan Ganz catheter via the jugular vein. Zero reference was set at the mid-chest. Cardiac output was measured by thermodilution technique and three values differing by <10% were averaged. Hemodynamic data were collected at baseline prior to further up-titration of vasoactive drugs, and about 3 hours after up-titration. As part of an ongoing institutional project to protocolize CS care^7^, this study has institutional approval (CARMS-17781) with identifiable data anonymized and patient consent was waived.

Hemodynamic response to up-titration of vasoactive drugs was quantified by the change in cardiac power output index (CPOI), which was calculated with the inclusion of right atrial pressure^8^:

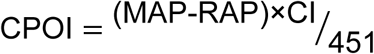

where MAP: mean arterial pressure, RAP, right atrial pressure and CI: cardiac index. Cardiac power output index about 3 hours after up-titration of inotropes and vasopressors (i.e. post-escalation CPOI) was used to assess hemodynamic response to treatment. ‘CPO responder’ was defined by a post-escalation CPOI of at least 0.28 W/m^2^, based on previous data^9^.

The vasoactive inotrope score (VIS), used to quantify the total doses of inotropes and vasopressors, was calculated as follows^10^:

VIS = dopamine (µg/kg/min) + dobutamine (µg/kg/min) + 100 × epinephrine (µg/kg/min) + 100 × norepinephrine (µg/kg/min) + 10 × milrinone (µg/kg/min) + 10,000 × vasopressin (units/kg/min)

### The ECMO-CS Cohort

The details and primary results of the Extracorporeal Membrane Oxygenation in the Therapy of Cardiogenic Shock (ECMO-CS) Trial have been published previously. In brief, this was a multicenter, randomized trial of immediate veno-arterial extracorporeal membrane oxygenation (VA-ECMO) or initial conservative management (no immediate VA-ECMO) in patients with rapidly deteriorating or severe cardiogenic shock, defined by echocardiographic, hemodynamic, and metabolic criteria. Importantly, in the early conservative arm, ECMO could be used in case of conservative therapy failure and further haemodynamic worsening with elevation in serum lactate.

There was no significant difference between treatment strategies in all-cause mortality and in composite endpoint of all-cause death, resuscitated cardiac arrest or implantation of another type of MCS^11,12^. A total of 117 patients were enrolled, 58 randomized to immediate ECMO and 59 to early conservative treatment. Baseline DBP were available for the whole cohort. Measurement of RAP was not required by the protocol; however, the values were recorded if available.

### Statistical analysis

Normality was assessed with the Kolmogorov-Smirnov test. Continuous variables were expressed as median (interquartile range, IQR) for non-normally distributed or mean ± SD for normally distributed variables. Comparisons were performed using Mann-Whitney’s (independent samples) or Wilcoxon signed rank test (paired samples) and t-tests or paired t-tests, for non-normally and normally distributed data respectively.

Multiple linear regression analysis was used to assess predictors of post-escalation CPOI, including baseline DPP, SVRI, cardiac index (CI), lactate and VIS. ‘CPO responder’ was defined by a post-escalation CPOI of at least 0.28 W/m^2^, and receiver operating characteristic curve was generated from the sensitivity and 1-specificity of baseline DPP for discriminating CPO responders from non-responders. The Youden index was used to identify the optimal cutoff point for baseline DPP, and the patients were characterized based on this optimal cutoff.

Supervised machine learning (logistic regression) was used to predict CPO responder. We tested 3 models based on the variables that were included in the model: Model 1 included baseline SVRI, VIS and lactate level; Model 2 included baseline DPP, VIS, CI and lactate level; and Model 3 is a simplified model that included only baseline DPP and CI. Firstly, train-test split is used to divide the data into two datasets (30: 70). The training dataset was used to train the model. Secondly, the testing dataset was used to check the model’s performance on ‘unseen’ dataset. The performance of the model was then assessed by the: (i) Precision (True Positive/(True Positive+False Positive)); (ii) Recall (True Positive/(True Positive+False Negative)), and (iii) the F1 score (2× (Precision×Recall)⁄(Precision+Recall)) metrics.

Time to death was analyzed using the Kaplan–Meier method and compared using the log-rank test. Hazard ratios (HRs) with corresponding 95% CIs were calculated using a Cox proportional hazard model with Efron approximation for tie holding.

All tests were two-sided with a p<0.05 considered statistically significant. All analyses were performed on Python (v3.13.1).

## RESULTS

### CS cohort

The baseline characteristics of the 93 patients with CS are presented in Table 1. Advanced heart failure was the cause of CS in most cases and deteriorating on inotropes (i.e. Society of Cardiovascular Angiography and Intervention (SCAI) stage D). The mean VIS at baseline was 11.2±3.9, with dobutamine or dopamine, milrinone or enoximone, epinephrine and norepinephrine used in 28, 66, 23, 83 patients respectively. Despite vasoactive drugs, baseline CI, DPP and CPOI were low at 1.62±0.24 L/min/m^2^, 37±7 mmHg and 0.18±0.04 W/m^2^, respectively. Baseline DPP correlated with heart rate (R^2^=-0.298, p=0.001) and SVRI (R^2^=0.567, p<0.001), but not CI [Supplementary Figure].

**Table 1:** Patient characteristics and hemodynamic data of the CS cohort (n=93)

| Parameter | Value |
| --- | --- |
| Age >50 years (n, %) | 51 (55%) |
| Males (n, %) | 67 (72) |
| BMI (kg/m<sup>2</sup>) | 26.9 (3.5) |
| Aetiology (n, %) |  |
| Acute myocardial infarction | 23 (25) |
| End-stage heart failure |  |
| - Ischemic | 38 (40) |
| - Non-ischemic | 23 (25) |
| Other | 9 (10) |
| NT-pro BNP (pg/ml) | 5032 (3019-9125) |
| Lactate (mmol/l) | 5.3 (4-6.3) |
| LVEF (%) | 15 (10-20) |
| Epinephrine (n, %) | 20 (22) |
| Norepinephrine (n, %) | 83 (89) |
| Vasoactive inotrope score | 11.2±3.9 |
| Systolic BP (mmHg) | 85 (6) |
| Diastolic BP (mmHg) | 54 (5) |
| Mean arterial BP (mmHg) | 67±5 |
| Heart rate (BPM) | 92 (85-105) |
| Right atrial pressure (mmHg) | 17±5.6 |
| PA systolic pressure (mmHg) | 57 (49-64) |
| PA diastolic pressure (mmHg) | 28 (24-31) |
| Mean PA pressure (mmHg) | 39 (35-44) |
| PA wedge pressure (mmHg) | 26 (23-30) |
| Cardiac index (L/min/m<sup>2</sup>) | 1.62±0.24 |
| Cardiac power output index (W/m<sup>2</sup>) | 0.18±0.04 |

BMI: body mass index; BP: blood pressure; LVEF: left ventricular ejection fraction; NT-proBNP: N-terminal B-natriuretic peptide; PA: pulmonary artery

Vasoactive drugs escalated by 13.3±2.6 to a VIS of 24.5±4.7. Escalation of vasoactive drugs was associated with increases in MAP (+4.1 (2.6-5.8) mmHg, p<0.001), heart rate (+4 (1-7) BPM, p<0.001), stroke volume index (+2 (1-3) ml/m^2^, p<0.001) and lower right atrial pressure (-1 (-2-1) mmHg, p=0.018) [Figure 1]. As a result, DPP, CI and CPOI increased to 41±8 mmHg, 1.89± 0.24 L/min/m^2^ and 0.23±0.06 W/m^2^ (all p<0.001).

**Figure 1:**
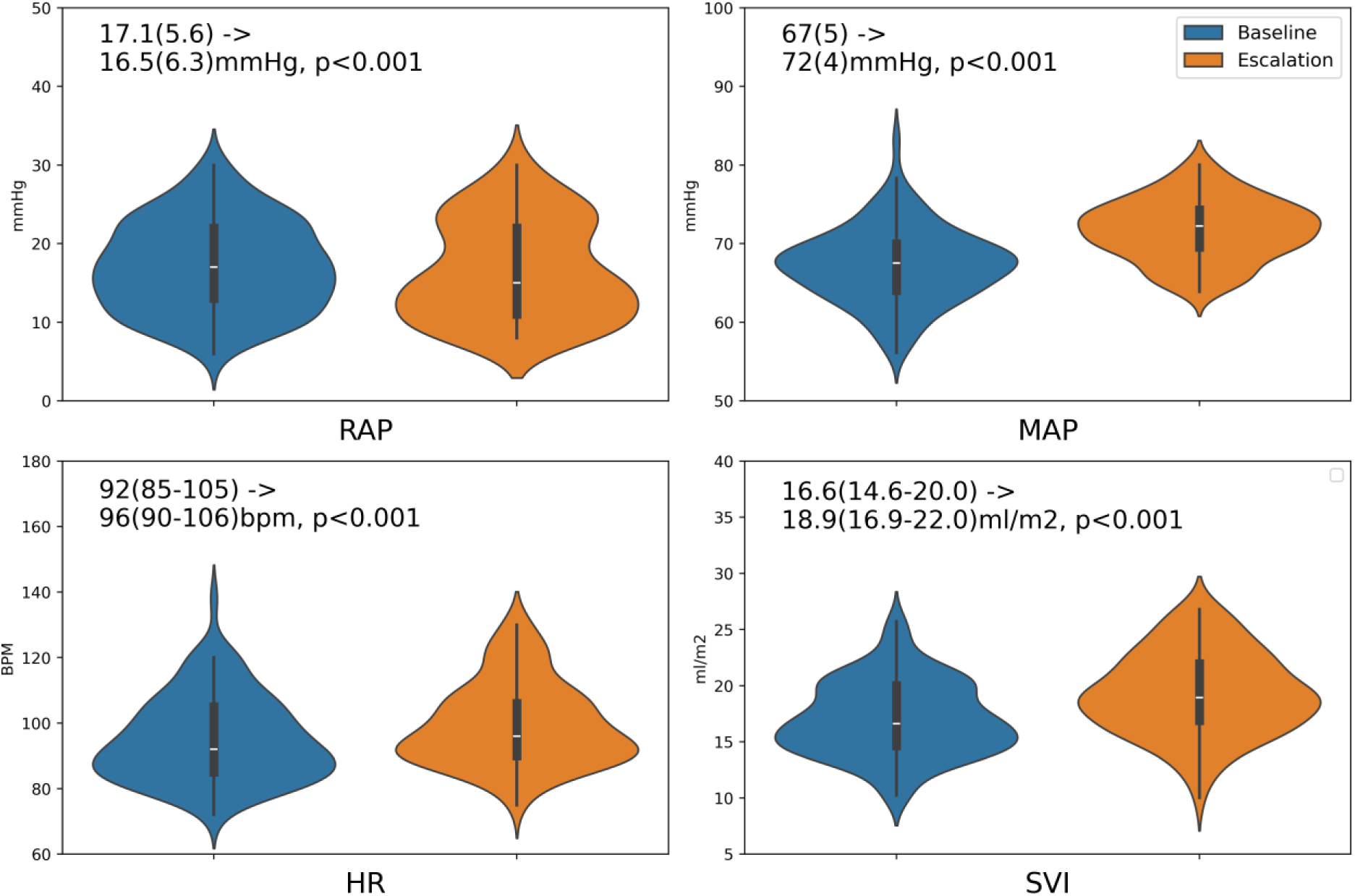
Violin plots of baseline and post-escalation hemodynamic parameters (RAP: right atrial pressure; MAP: mean arterial pressure; HR: heart rate; SVI: stroke volume index).

Post-escalation CPOI was related to baseline CI and DPP [Figure 2] on multiple linear regression analysis [Supplementary Table]. Of the 93 patients, 35 patients achieved post-escalation CPOI of ≥0.28 W/m^2^ (‘CPO responders’). None of the CPO responders had baseline DPP of <36mmHg, CI of <1.3L/min/m^2^ or CPOI of <0.158W/m^2^. The maximum increase in CI and CPOI were 0.69 L/min/m^2^ and 0.123W/m^2^, respectively.

**Figure 2:**
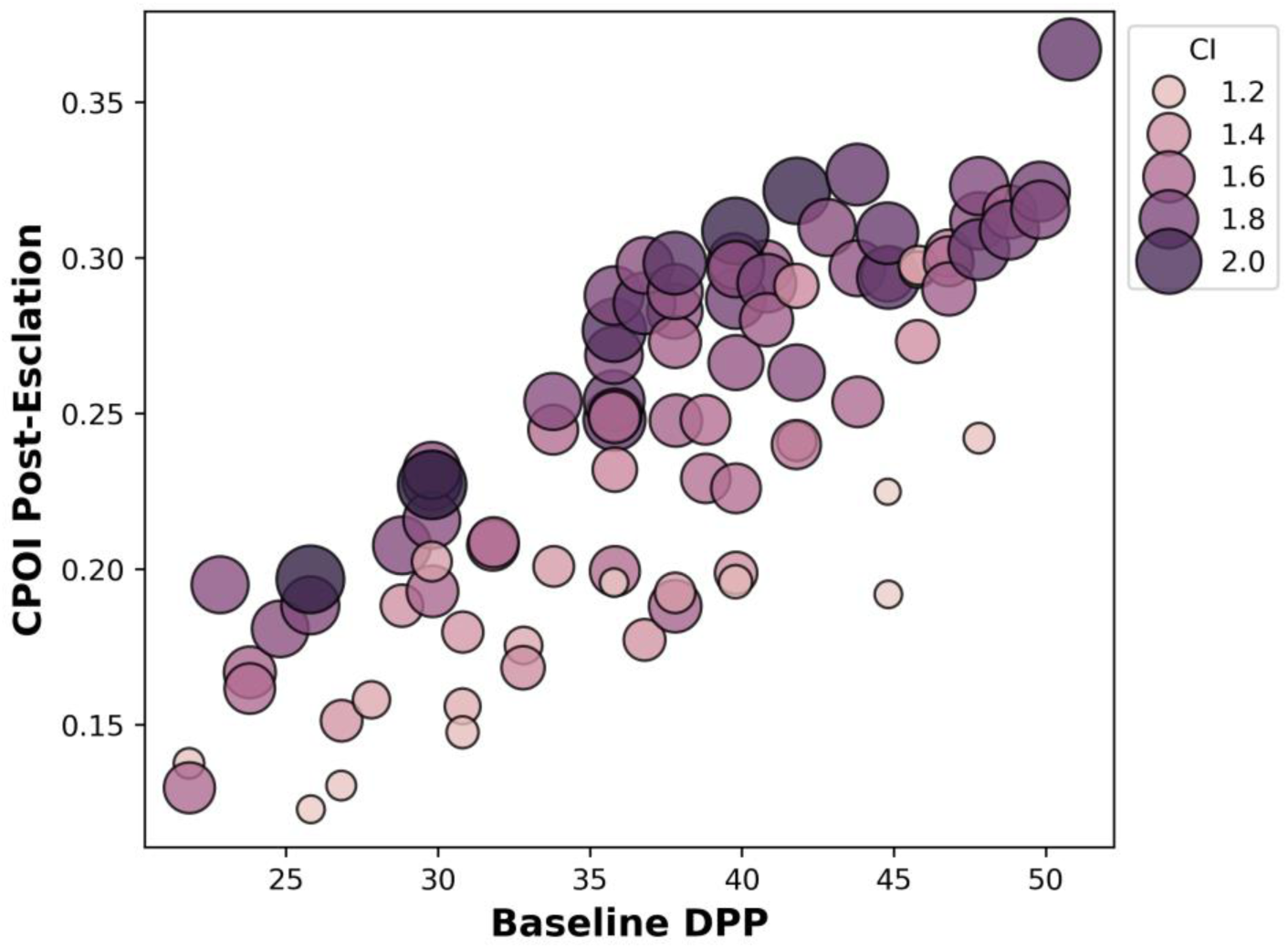
Post-escalation CPOI is associated with baseline DPP and CI. Baseline DPP and CI were higher in patients who achieved higher post-escalation CPOI.

From ROC curves, baseline DPP discriminated CPO responders from non-responders well compared to SVRI [Supplementary Figure]. The optimal baseline DPP cutoff of 37mmHg based on Youden Index [Supplementary Table]. We divided the cohort into two subgroups based on this baseline DPP cutoff: a low DPP subgroup with baseline DPP <37mmHg (n=43) and a second subgroup with baseline DPP ≥37mmHg (n=50). The low DPP subgroup had significantly lower post-escalation CPOI and lactate clearance despite greater increase in VIS [Figure 3]. Indeed, despite lower lactate at baseline, blood lactate was higher at 3 hours in the low compared to the DPP≥37mmHg subgroup [Supplementary Figure]. Using supervised machine learning (logistic regression), the simplified Model 3 that included only baseline DPP and CI outperformed Model 1 (baseline SVRI, VIS and lactate) and Model 2 (baseline DPP, VIS and lactate), with highest precision (0.83, vs 0.64 and 0.82), recall (0.83 vs 0.57 and 0.81) and F1 score (0.79 vs 0.60 and 0.79).

**Figure 3:**
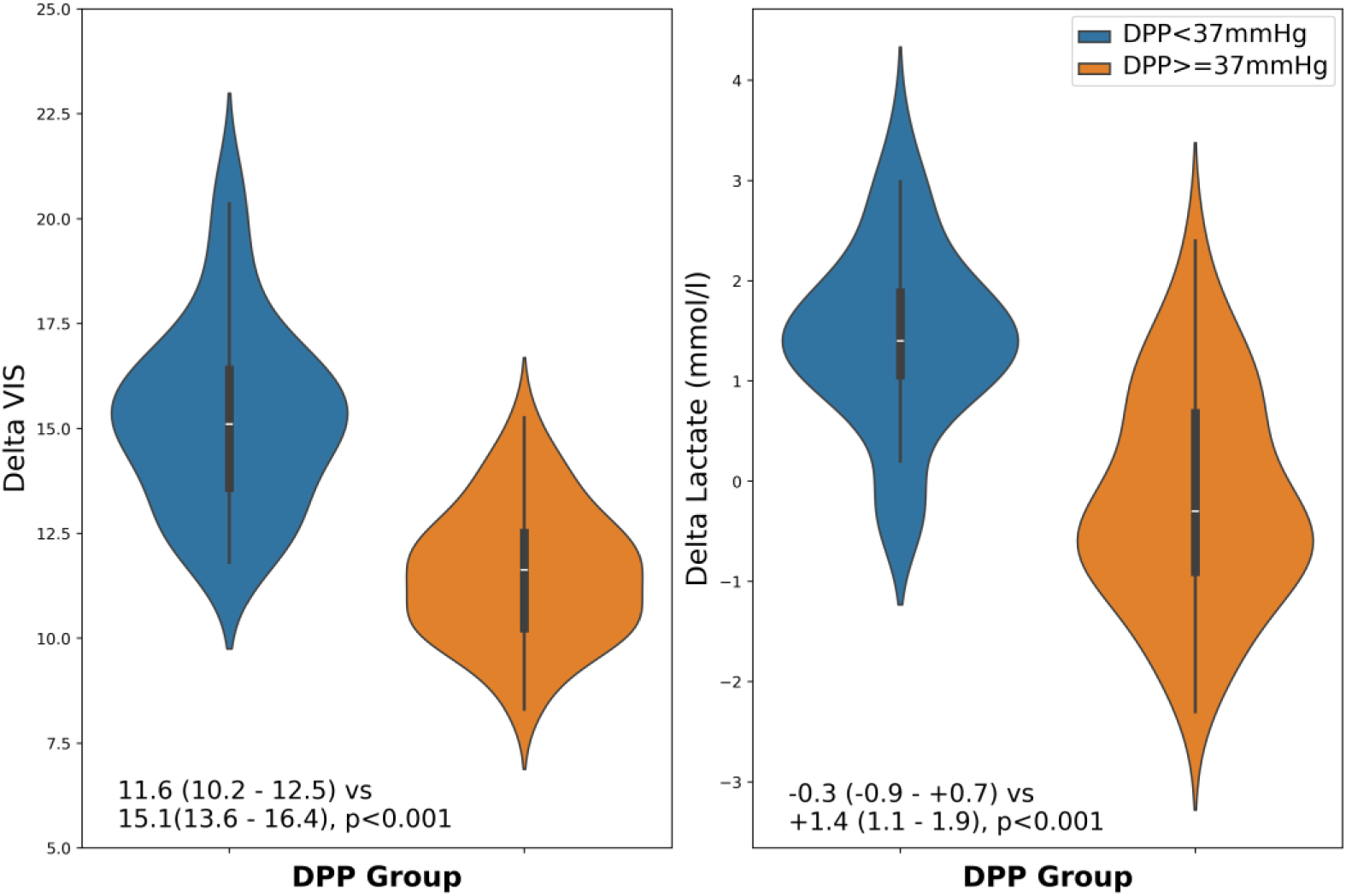
Violin plots of changes in VIS and lactate (post-escalation minus pre-escalation) in the DPP<37mmHg compared to the DPP≥37mmHg subgroup. The DPP≥37mmHg subgroup had a smaller increase in VIS and greater reduction in blood lactate at about 3 hours compared to the DPP<37mmHg subgroup.

### ECMO-CS cohort

Baseline DPP values were available in 63 of 117 randomized patients. The same DPP cutoff of 37mmHg was applied to dichotomize the ECMO-CS cohort into two subgroups. The immediate VA ECMO subgroup with DPP ˂37 mmHg had lower VIS compared to the early conservatively treated subgroup. The subgroups with DPP ≥37 mmHg had higher blood pressure and lower lactate in comparison to patients with DPP ˂37 mmHg. The characteristics of the patients in the ECMO-CS trial, dichotomized by this DPP cutoff of 37mmHg are shown in Table 2.

**Table 2:** Baseline characteristics of the ECMO-CS cohort by DPP cutoff of 37mmHg.

|  | **DPP < 37mmHg** |  | **DPP ≥ 37 mmHg** |  |
| --- | --- | --- | --- | --- |
|  | **VA ECMO** | **Conservative** | **VA ECMO** | **Conservative** |
| **Number of patients** | 12 | 16 | 18 | 17 |
| **Age (years)** | 71 (61-74) | 67 (62-74) | 68 (61-73) | 66 (59-73) |
| **Female (%)** | 50 | 19 | 18 | 35 |
| **BMI (kg/m<sup>2</sup>)** | 27 (25-29) | 29 (26-31) | 26 (23-30) | 28 (25-31) |
| **STEMI (%)** | 42 | 44 | 71 | 47 |
| **PCI (%)** | 42 | 50 | 71 | 53 |
| **Lactate (mmol/l)** | 6.0 (4.0-7.8) | 6.7 (3.4-11.0) | 4.6 (3.2-8.0) | 3.6 (3.1-5.3) |
| **Systolic BP (mmHg)** | 80 (70-83) | 80 (70-90) | 90 (85-100) | 100 (80-105) |
| **Diastolic BP (mmHg)** | 45 (40-50) | 45 (40-50) | 60 (55-60) | 60 (60-60) |
| **MAP (mmHg)** | 60 (50-61) | 54 (46-61) | 70 (65-70) | 70 (65-80) |
| **CVP (mmHg)** | 16 (12-21) | 16 (15-19) | 13 (9-17) | 15 (12-18) |
| **VIS** | 54 (31-102) | 131 (52-198) | 27 (3-42) | 45 (24-73) |
| DPP, diastolic perfusion pressure; BMI, body mass index; STEMI, ST-elevation myocardial infarction; PCI, percutaneous coronary intervention; BP, blood pressure; MAP, mean arterial pressure; CVP, central venous pressure; VIS, vasoactive inotropic score |  |  |  |  |

Clinical outcomes were related to baseline DPP. In the immediate VA ECMO arm, patients with DPP ˂37mmHg had significantly lower one-year all-cause mortality (HR 0.37, 95% CI 0.14-0.97, P = 0.044), but in the subgroup with DPP ≥37mmHg, immediate VA ECMO compared to early conservative arm did not increase chance of survival (Figure 4).

**Figure 4:**
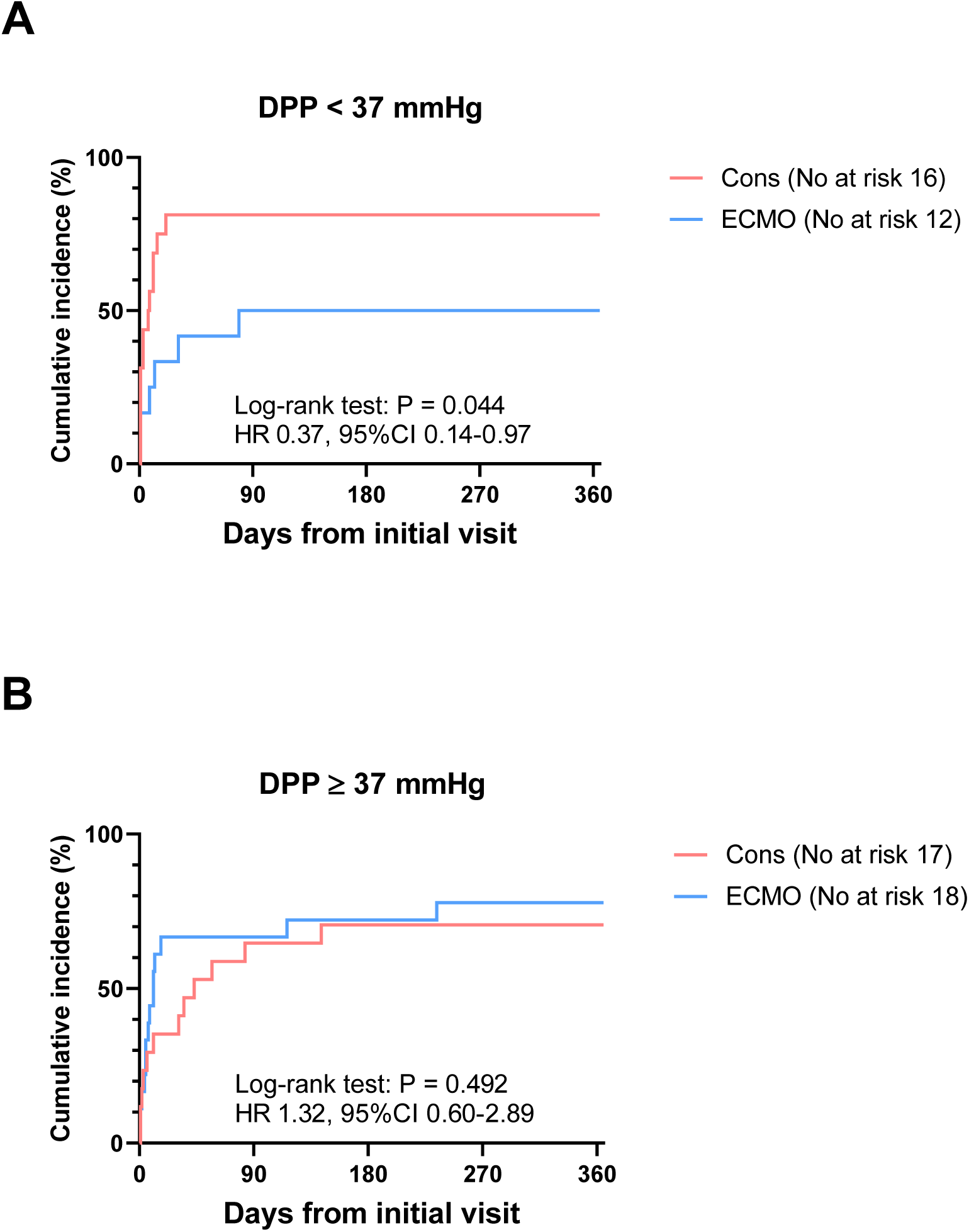
Cumulative incidence of all-cause mortality in the ECMO CS cohort, divided into two groups based on pre-ECMO DPP of <37mmHg (Panel A) or ≥37mmHg (Panel B).

In the early conservative arm, 11 of 16 (69%) patients with DPP ˂37 mmHg experienced hemodynamic worsening requiring VA-ECMO and all 3 survivors were on VA ECMO. In contrast, in early conservatively treated patients with DPP ≥37 mmHg, only 6 of 17 (35%) patients required “bailout” ECMO and 4 of 5 survivors were without ECMO.

## DISCUSSION

Understanding the hemodynamic response to escalation of vasoactive drugs is central to addressing the “clinician’s dilemma”. In the first part of this study of patients with SCAI D CS, we have shown that (i) baseline CI and DPP predicted CPOI response to escalation of vasoactive drugs; (ii) patients with baseline DPP ≥37mmHg were more likely to achieve CPOI of ≥0.28 W/m^2^ (‘CPO responders’) and greater lactate clearance; and (iii) applying this DPP cutoff to a cohort of patients in the ECMO-CS trial, we found that patients with DPP˂37mmHg had higher survival with immediate VA ECMO compared to early conservative management, but not in the subgroup with DPP≥37mmHg.

The terms vasoplegia, distributive/ vasodilatory shock and vascular failure are often used interchangeably to infer a state of pathological dilatation (or loss or vasomotor tone) of resistance arterioles. For the sake of consistency, we have used the term abnormal vasodilation. The phenotype of CS with concomitant abnormal vasodilatation has been termed ‘mixed shock’ and a conceptual model based on the presumed primary insult has been proposed. However, there is no agreed definition for abnormal vasodilatation or ‘mixed shock’, although most studies have used low systemic vascular resistance^13^. Based on the physiological considerations [Supplementary Data], we have proposed DPP as a measure of vasodilatation. In this study, DPP, but not SVRI was associated with CPOI response, a finding that is consistent with our previous study of hemodynamic instability post-heart transplant.

The physiology of cardiac output regulation may explain our finding that baseline CI and DPP predicted CPOI response to escalation of vasoactive drugs. Cardiac stroke volume is preload-dependent and sensitive to afterload. Stroke volume is expected to be greater at the same preload and contractility in the face of low afterload. As such, stroke volume and CI should be higher in patients with CS and concomitant abnormal vasodilatation (i.e. ‘mixed shock’) compared to patients with CS without concomitant abnormal vasodilatation, if contractility and preload were comparable. Thus, the lower CI in patients with CS and abnormal vasodilatation implies more severe contractile dysfunction, as preload appeared to be comparable. The more severe contractile dysfunction may limit response to inotropes and may be more sensitive to increases in afterload. The implication is that vasoconstriction would further limit stroke volume response to inotropes, especially at the limit of recruitable preload. In addition, low afterload produces a ‘low gain’ system such that every unit increase in stroke volume would produce a smaller ‘gain’ in blood pressure^14^. A ‘low gain’ system limits blood pressure response to increased stroke volume from inotropes. Thus, the poorer CPOI response to escalation of vasoactive drugs may reflect a combination of more severe contractile dysfunction that limits stroke volume response to escalation of inotropes and greater afterload sensitivity, and a low system ‘gain’ due to abnormal vasodilatation.

Our study has direct clinical application. Firstly, DPP may be used to identify patients who are less likely to response to up-titration of vasoactive drugs and guide timely deployment of MCS. The optimal baseline DPP cutoff of 37mmHg from the Youden Index was based on conventional assumption of equivalence between sensitivity and specificity. However, the sensitivity and specificity thresholds may be altered depending on prioritization of the risk of delayed MCS vs the risks of ‘excess’ MCS use. For example, there were no CPO responders in patients with DPP <36mmHg. Therefore, with this more stringent cutoff of <36mmHg, there will be no ‘excess’ MCS use but waiting for the DPP to drop below <36mmHg would delay the deployment of MCS, delays that could result in failure of MCS therapy. In contrast, if the objective is to avoid delays in MCS therapy, a cutoff of 40mmHg may be preferred, at the expense of an ‘excess’ MCS use.

We further examined this DPP cutoff of 37mmHg in a cohort of patients from the ECMO-CS trial to externally validate this parameter. ‘Immediate’ VA ECMO in patients with DPP ˂37mmHg was associated with significantly higher probability of survival (but not in the subgroup with DPP ≥37mmHg). Crucially, there were no survivors without VA ECMO in patients with DPP <37mmHg in the early conservative arm of the ECMO-CS trial. It is possible that low DPP identified a subgroup who had poor response to vasoactive drugs and thus, might have more to benefit from early MCS. The limitations of post hoc analysis notwithstanding, this finding from the ECMO-CS trial cohort provides validation for this parameter and supports the use of DPP in guiding the use of MCS.

Secondly, none of the CPO responders had baseline DPP of <36mmHg, CI of <1.3L/min/m^2^ or CPOI of <0.158W/m^2^, with baseline vasoactive drug treatment at VIS of 13.3±2.6. These criteria may be used to identify a high-risk cohort in whom early MCS may be indicated. For example, a clinician facing a deteriorating patient with CS on a combination of vasoactive drugs at VIS of 15 may opt for MCS if the DPP and CI were 35mmHg and 1.3L/min/m^2^ respectively, as the hemodynamic goals may not be achievable with further escalation of vasoactive drugs. Similarly, increasing VIS to 24.5±4.7 produced a maximum increase in CI of 0.69 L/min/m^2^. On this basis, patients with baseline CI of <1.5L/min/m^2^ are unlikely to achieve CI of 2.2L/min/m^2^, if this is the CI required to achieve adequate oxygen delivery as part of goal-directed therapy in CS. In this regard, the use of pulmonary artery catheter to monitor CI and CPOI may be useful to guide timely deployment of MCS in patients with deteriorating CS.

### Study limitations

This study has several limitations. Firstly, our CS cohort is relatively small and derived from a single center, and thus at risk of bias. The observational nature is also inherently susceptible to biases, including biases associated with selection of patients. Secondly, our analysis of the ECMO-CS trial is at risk of bias as with any post hoc analysis of clinical trials. Therefore, our findings should be considered hypothesis-generating and should be confirmed in future studies. Thirdly, the choice and titration of vasoactive drugs were left to the discretion of the clinicians, guided by hemodynamic assessment from pulmonary artery catheterization. It is unclear if a more protocolized strategy for titration of vasoactive drugs would have produced greater hemodynamic response. Similarly, it is not clear if even greater increase in VIS could have achieved the CPOI target and result in greater lactate clearance. Finally, we did not directly compare up-titration of vasoactive drugs against the use of MCS. The benefit of MCS over escalation of vasoactive drugs in patients with low DPP needs to be confirmed.

## CONCLUSION

Low baseline DPP and CI were associated with poor CPOI response to escalation of inotropes and vasopressors. Baseline DPP <37mmHg was associated with lower CPOI response and lactate clearance. In a post hoc analysis of the ECMO-CS trial, ‘immediate’ VA ECMO in patients with DPP ˂37mmHg was associated with significantly higher probability of survival and there were no survivors without VA ECMO in patients with DPP <37mmHg in the early conservative arm of the ECMO-CS trial.

## Data Availability

Some data may be made available upon request

## Abbreviations

CI: cardiac index
CPOI: cardiac power output index
CS: cardiogenic shock
DBP: diastolic blood pressure
DPP: diastolic perfusion pressure
ECMO: extracorporeal membrane oxygenation
HR: heart rate
MAP: mean arterial pressure
MCS: mechanical circulatory support
SBP: systolic blood pressure
SVI: stroke volume index
SVRI: systemic vascular resistance index
VIS: vasoactive inotrope score

